# Evaluate the risk in conventional IVF frozen human blastocysts undergoing PGT using a new quantification method for parental contamination testing (qPCT)

**DOI:** 10.1101/2022.01.23.22269520

**Authors:** Yunqiao Dong, Dun Liu, Yangyun Zou, Cheng Wan, Chuangqi Chen, Mei Dong, Yuqiang Huang, Cuiyu Huang, Huinan Weng, Xiulan Zhu, Fang Wang, Shujing Jiao, Na Liu, Sijia Lu, Xiqian Zhang, Fenghua Liu

**Affiliations:** Reproductive Medical Center, Guangdong Women and Children Hospital, Guangzhou Medical University, Guangzhou, 511400, Guangdong Province, China; Yikon Genomics Company, Ltd., Shanghai 201499, China

**Keywords:** Quantitative parental contamination testing, IVF, PGT, biopsy, human blastocyst

## Abstract

**Objective:** To establish a method to assess risks associated with preimplantation genetic testing (PGT) in embryos simultaneously with adhered sperm and cumulus cells.

**Design:** A prospective pilot study.

**Setting:** University teaching hospital.

**Patient(s):** 120 frozen blastocysts that could be biopsied from 34 patients who had experienced repeated implantation failure or abortion due to chromosomal abnormalities after embryos transfer in prior routine IVF cycles.

**Intervention(s):** Chromosome screening and parental DNA contamination testing was performed in the surplus frozen IVF blastocysts from 34 patients.

**Main Outcome Measure(s):** Parental DNA contamination rate and euploidy rate in biopsied blastocysts.

**Result(s):** A new quantification method for parental contamination testing (qPCT) in single-cell whole-genome amplification (WGA) products based on allelic ratio analysis was established and validated in an artificial model by comparing 22 results obtained before and after adding different numbers of sperm and cumulus cells to biopsied TE cells. The results of the prospective clinical study of qPCT-PGT-A showed that the maternal contamination rate was 0.83% (1/120) and the risk of paternal contamination was negligible. The euploidy rate in these blastocysts was 47.50% (57/120), and 21 frozen embryo transfer (FET) cycles resulted in ten ongoing clinical pregnancies and four healthy births.

**Conclusion(s):** The evidence we provide in the study shows a low risk of PGT in embryos simultaneously with adhered sperm and cumulus cells. The qPCT assay can be used to detect the risk of potential contamination and ensure the accuracy of PGT results, thereby improving the clinical outcome of IVF.

**Capsule:** During PGT for human frozen conventional IVF embryos, paternal source pollution is negligible, while maternal pollution can not be ignored. qPCT method can effectively detect the parent DNA contamination in WGA products of biopsied TE cells.

## Introduction

For preimplantation genetic testing (PGT) to be clinically applicable, intracytoplasmic sperm injection (ICSI) is usually required, rather than conventional in vitro fertilization (IVF) insemination method, owing to potential parental contamination from sperm and cumulus cells attached to the zona pellucida during conventional IVF (1-6). However, in many available studies addressing non-male factor infertility, ICSI has not been proven to be beneficial for clinical outcomes, and the risk of chromosomal defects is increased in ICSI offspring (7-12). Since sperm DNA fails to amplify under the conditions used for trophectoderm (TE) biopsy samples, and cumulus cells will be deliberately removed clearly in fresh IVF-PGT-A, the use of conventional IVF has been preliminarily explored in fresh genetic testing for aneuploidies (PGT-A) cycles in recent years (13-15). However, if without an intention to perform PGT in the beginning, there are many sperm and cumulus cells exist simultaneously in the zona pellucida of conventional IVF frozen blastocysts. Little is known about the maternal contamination risk of PGT in these embryos and whether it adversely affects the accuracy of results in conventional IVF-PGT. Recently, a study shows that 12.5% maternal DNA contamination was observed in blastocyst media, indicating that potential maternal contamination should not be neglected (16).

Currently, the clinical need for PGT-A in frozen conventional IVF embryos is urgent. For patients who have experienced repeated implantation failure or abortion due to chromosomal abnormalities in previous embryo transfer cycles, the biopsy of the existing frozen embryos is not only necessary but also cost-effective. However, there are many cumulus cells and sperm in the zona pellucida especially from conventional IVF embryos, especially when there is no PGT expectation in advance. With current methods, parental contamination in biopsy cells cannot be detected accurately. Therefore, we verified whether PGT-A is feasible for embryos with potential parental contamination.

To address this issue, we developed a parental contamination detection method for biopsy cells which could exclude contaminated embryos and ensure the accuracy of PGT results. Thus far, several approaches using single nucleotide polymorphism (SNP) genotyping information or short tandem repeat (STR) markers to trace maternal genetic materials from fetal or miscarriage specimens have been reported (17-19). However, such approaches require sufficient DNA for genetic testing. Esteki and his collaborators proposed a concurrent whole-genome haplotyping approach referred to as haplarithmisis to resolve the parental origin of copy number variations (CNVs) in single cells, but the method is easily affected by low-quality amplification effects, such as high allele dropout (ADO) (20). To the best of our knowledge, no previous studies have reported parental contamination detection in single-cell whole-genome amplification (WGA) products from TE biopsied cells.

In this study, we first developed a new quantification method for parental contamination testing (referred to as qPCT) based on allelic ratios obtained from SNP genotypes that fully considers amplification bias, including preferential amplification or ADO effects. The qPCT assay was validated by comparing results before and after the artificial addition of sperm and cumulus cells to TE cells from discarded aneuploidy blastocysts. We then recruited 120 frozen embryos from 34 couples, with advanced age, experienced repeated implantation failure or abortion due to chromosome abnormalities, who had surplus frozen embryos from previous conventional IVF cycles available for biopsy. By using next-generation sequencing (NGS) with our qPCT method, we selected euploidy embryos with no parental contamination for transfer, resulting in four healthy live birth and ten ongoing clinical pregnancies. The method has the potential for application in much wider chromosome screening in clinical IVF due to increasing clinical needs and the limitations of the existing methods.

## Materials and Methods

### Preparation of embryos for the study

To establish the qPCT method, 30 reference samples, which are WGA products with normal PGT-A results and no parental contamination were collected, and to perform the simulated contamination test, six discarded aneuploidy embryos were collected. We conducted a prospective clinical study with patients recruited from January 2020 to April 2021. A total of 34 couples with indications for PGT-A due to advanced maternal age, repeated implantation failures or recurrent abortion who had surplus blastocysts from prior conventional IVF available for biopsy were recruited regardless of their prognosis, age, or diagnosis. The couples had no karyotype abnormalities. Approval for this study was obtained from the Ethics Committee of Guangdong Women and Children Hospital (Research Ethics Committee No. 202101109). All these samples were from Reproductive Medical Center of Guangdong Women and Children Hospital and all of the patients signed the informed consent form before any study-specific procedures were performed. The whole protocol described below is summarized in Figure. 3A. The clinical details of 34 couples are summarized in Table 2.

### Artificial addition of sperm and cumulus cells to TE cells

To establish an artificial model for detecting parental cells contamination in biopsied TE cells, frozen-thawed sperm and cumulus cell was isolated, aspirated and put into the pipette with biopsy cells. To simulate the freezing process of sperm on the zona pellucida, the method involving the zona pellucida combined with sperm was used (Supplementary, Figure. S1).

### Blastocyst biopsy

Frozen stage 5 or 6 blastocysts were thawed in advance and cultured 2-4 h prior to biopsy, whereas frozen day 3 embryos were thawed 2–3 days in advance. Every blastocyst that reached the blastocyst stage and fulfilled the biopsy criteria was biopsied. Biopsy was performed in 10-μl drops of G-MOPS-PLUS, the blastocyst was fixed and positioned with a clear view of the inner cell mass (ICM) at 9-12 o’clock, and the hatched TE cells were aspirated into the biopsy pipette, followed by three laser pulses of 2.0 ms (ZYLOS-tk^®^, Hamilton Thorne, MA, USA) to loosen cell connections and the application of the mechanical ‘flicking’ method to cut away the TE cells using the biopsy pipette and holding pipette. When a blastocyst was not hatched, the zona pellucida was perforated by a laser for 2.0 ms, after which the collapse of the blastocyst was induced before biopsy. TE cells were washed and placed in 0.2-ml PCR tubes with 1–2.0 μl PBS and stored at −20°C until further processing. Approximately 10 biopsy cells from one discarded aneuploidy blastocyst were artificially mixed with sperm or cumulus cells and 4–6 TE cells were biopsied for clinical application of qPCT-PGT. All embryos were cryopreserved by vitrification (Life Global^®^, USA) within 1 h after biopsy and stored at -196°C.

### Whole-genome amplification of sperm, cumulus and TE cells

Single-cell WGA were performed in 22 samples of sperm cells (20 cells per sample) and three samples of cumulus cells (one cell per sample) under different conditions used for TE biopsy samples (Figure. 1A and 1B). The multiple annealing and loop-based amplification cycles (MALBAC)-based single-cell WGA kit (ChromSwiftTM, Cat. No. KT110700324, Yikon Genomics Ltd, Suzhou, China), the PicoPLEX single-cell WGA kit (Rubicon Genomics, Ann Arbor, USA) and the multiple displacement amplification (MDA)-based single-cell WGA Kit (Cat. No. 150343, Qiagen, USA) were utilized to amplify DNA from different types of cells according to the manufacturer’s instructions.

**Figure 1.**
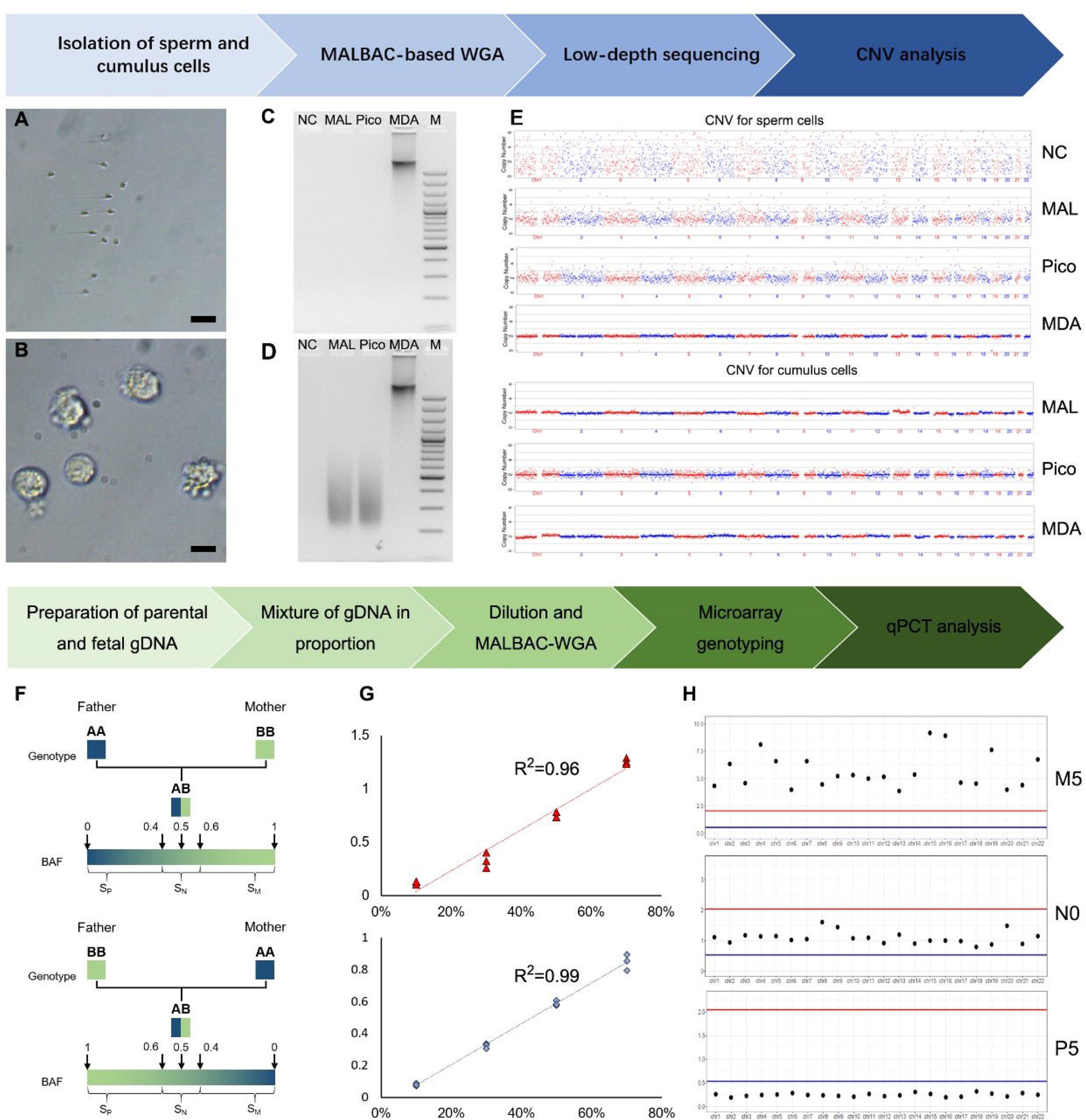
Concept validation and establishment of qPCT. **A and B:** Isolated sperm and cumulus cells. Bar = 10 μm. **C and D:** Electrophoresis for WGA products of sperm and cumulus cells. M: 100 bp DNA marker; NC: negative control; C-MAL: 20 sperm cells amplified by MALBAC; C-Pico: 20 sperm cells amplified by Picoplex; C-MDA: 20 sperm cells amplified by MDA. D-MAL: single cumulus cell amplified by MALBAC; D-Pico: single cumulus cell amplified by Picoplex; D-MDA: single cumulus cell amplified by MDA. **E:** CNV-seq results of WGA products amplified sperm and cumulus cells using MALBAC/Picoplex/MDA. Upper: sperm cells; lower: single cumulus cell; **NC:** negative control. **F:** Schematic of assignment of SNP sites for paternal/maternal bias. **S**_**M**_: SNP site for maternal bias; S_P_: SNP site for paternal bias. **G:** Standard curve of qPCT established by artificially mixed parental and fetal gDNA. **X-**axis: ratio of added maternal/paternal gDNA; Y-axis: POB value (log_10_); Red triangle: maternal contamination samples; Blue rhombus: paternal contamination samples. **H:** qPCT results for artificially contaminated gDNA samples. X-axis: chromosome ID; Y-axis: POB value; M5: 50% maternal contamination sample (maternal DNA : fetal DNA = 1:1); N0: no contamination sample; P5: 50% paternal contamination sample (paternal DNA : fetal DNA = 1:1).

### Determination of blastocyst ploidy status by NGS

To analyze the ploidy status of blastocysts, the amplified DNA of TE samples was sequenced using a NextSeq 550 sequencer (Cat No. SY-415-1002, Illumina, Inc., USA) with a single-ended read length of 55 bp. Approximately 2 million raw reads were generated for each TE sample. Genome-wide CNVs were analyzed to determine the euploidy or aneuploidy status of each embryo. Aneuploid embryos were diagnosed when the size of CNVs was greater than 4 Mb and the extent of mosaicism was above 30%.

### Genotyping assay of blastocysts and parental genomes

To determine the genotype of each blastocyst and parental genome, the Infinium Asian Screening Array (ASA) bead chip (Cat No. 20016317, Illumina, Inc., USA) and the iScan system (Cat No. SY-101-1001, Illumina, Inc., USA) were utilized. Parental genomic DNA was extracted from the peripheral blood of the parents of each blastocyst. Genomic DNA, along with the amplified DNA of TE samples, was linearly amplified, fragmented, precipitated and hybridized according to the manufacturer’s instructions. Signal scanning was performed with the iScan system. Iaap-cli gencall algorithm (Version 1.1.0, Illumina, Inc., USA) was applied to analyze the genotype of each sample, and the BAF and log R ratio (LRR) values of all SNPs were generated simultaneously.

### Determination of parental contamination by BAF analysis

As shown in (Figure. 1F), theoretically, the BAFs of genotypes AA, AB and BB should be approximately 0, 0.5 and 1, respectively. For a SNP with paternal AA and maternal BB genotypes or paternal BB and maternal AA genotypes, the genotype of the embryo is genetically expected to be heterozygous AB, with a BAF of approximately 0.5. Deviation from a BAF of 0.5 for the embryo may be caused by parental contamination, DNA copy number aberrations or allelic amplification bias and errors (e.g. allele dropout or preferential amplification of one allele) resulting from single-cell WGA technology. Selecting SNP j with homozygous genotypes but different alleles between the father and mother, maternally and paternally biased SNPs in an embryo were defined according to a BAF ≥0.6 (S_M_, SNP site for maternal bias) and BAF ≤0.4 (S_P_, SNP site for paternal bias) given maternal and paternal genotypes of BB and AA, respectively; for maternal and paternal genotypes of AA and BB, maternally and paternally biased SNPs were determined according to a BAF ≤0.4 (S_M_) and BAF ≥0.6 (S_P_) (Figure. 1F). Relative parental origin bias (POB) statistics were thus constructed and formulated for each chromosome, i, as follows:

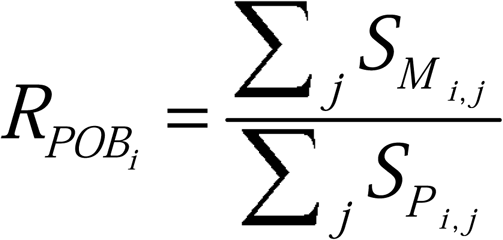

Based on reference samples from euploid embryos without parental contamination, a positively skewed normal distribution of 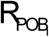 with a mean value of 1.14 was obtained (Figure. S1B). Here, a tested sample with an 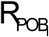 value significantly greater than 1.14 is indicated to show maternal origin bias, implying maternal contamination (most chromosomes with numbers ≥15 share the same pattern), or chromosome-level maternal-origin DNA replication, or uniparental disomy, or paternal-origin DNA deletion with a *P*-value < 0.05 (Mean+3*SD); on the contrary, a tested sample with an 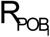 value significantly less than 1.14 is indicated to show paternal-origin bias, with the possibility of paternal contamination (most chromosomes with numbers ≥15 share the same pattern), or chromosome-level paternal-origin DNA replication, or uniparental disomy, or maternal-origin DNA deletion with a *P*-value <0.05 (Mean-2*SD, standard deviation). *R*_*POB*_ statistics can to some degree eliminate allelic amplification bias from WGA.

### Preparation and analyzing of artificially contaminated gDNA

To address the issue of quantification of parental contamination, we establish a standard curve, which represents the correlation between the POB value and the ratio of parental contamination. Parents genomic DNA was extracted from peripheral blood samples, while fetal genomic DNA was extracted from tissue of spontaneous abortion. A commercial genomic DNA extraction kit (DNeasy Blood & Tissue Kit, Cat. No. 69504, Qiagen, USA) was used according to the manufacturer’s instructions. Paternal/fet al and maternal/fet al DNA was mixed in different proportion (Figure. 1G). The artificially contaminated DNA was then diluted to 50pg/μL, and amplified by using a ChromInst Single cell WGA kit (Cat. No. Yikon Genomics Ltd, Suzhou, China) according to the manufacturer’s instructions. Finally, the amplified DNA was analyzed by using the Infinium Asian Screening Array (ASA) bead chip (Cat No. 20016317. Illumina, Inc. USA) and the iScan system (Cat No. SY-101-1001. Illumina, Inc. USA) to obtain the BAF value of each SNP loci. The R_POB_ was calculated to establish the standard curve of the ratio of parental contamination.

### Measurement of clinical outcomes

Biochemical was defined as β-HCG > 5 U/L detected 12 days after blastocyst transfer, but no gestational sac was found by transvaginal ultrasound. Negative was defined as β-HCG <5 U/L detected 12 days after blastocyst transfer. Clinical pregnancy was defined by fetal cardiac activity. Non-invasive prenatal testing (NIPT) was performed at 12−18 weeks of gestation.

## Results

### WGA of sperm and cumulus cells

We performed WGA and copy number variation sequencing (CNV-seq) in 22 samples of frozen-thawed sperm cells and three samples of frozen-thawed cumulus cells (Figure 1A and 1B). As shown in Figure. 1C, no DNA smears were observed in the MALBAC-WGA and Picoplex-WGA products of the sperm samples, while positive results were obtained from the MDA-WGA products. However, cumulus cell DNA can be easily amplified by the three methods (Figure 1D). The MALBAC-CNV-seq and Picoplex-CNV-seq of sperm samples also failed to generate qualified results, showing low genome coverage of 0.31% to 1.08% and relatively high coefficients of variation (CV) ranging from 23% to 50% (Figure 1E). However, three cumulus cell samples presented high-quality CNV-seq results, showing normal karyotypes of the three samples (Figure 1D and 1E). These results are partially in accordance with previous studies (15), indicating the difficulty of amplifying sperm DNA.

### Establishment of qPCT

To establish the qPCT method, we first collected 30 reference samples from euploid embryos without parental contamination, which were defined as embryos with normal ploidy and XY chromosomes. The POB values of these reference samples were calculated based on the B allele frequency (BAF) of selected SNP loci (Figure. 1F and Supplementary Figure. S2) approximately followed a positively skewed normal distribution, with a mean (Mean) of 1.14 and standard deviation (SD) of 0.30. Therefore, the POB range for non-contaminating euploid embryos was defined as 0.54 to 2.04 (Mean -2*SD to Mean + 3*SD), and a qualitative standard of parental contamination was established. To further address the issue of the quantification of parental contamination, gDNA of tissue samples from spontaneous abortions and blood samples from parents was mixed in specific proportions. The mixed gDNA was diluted to an approximate concentration of 50 pg/μL and then amplified, sequenced and analyzed in accordance with the PGT-A procedure. Genotyping assays were also performed. As a result, the POB values of the artificially added contaminant fet al gDNA showed a positive correlation with the proportion of mixed parental gDNA (Figure 1G and 1H). The relative quantification of parental contamination could be performed by using the standard curve.

### Detection of contamination in artificially contaminated TE samples

To verify the accuracy and repeatability of qPCT in detecting parental contamination in biopsy cells, we added gDNA from the peripheral blood of parents to the WGA products and found that the detection rate was 100% (10/10) (Supplementary Figure S3). However, no significant contamination was detected in artificially contaminated TE samples containing approximately 10 biopsy cells and 1 cumulus cell (Table 1).

**Table 1.**
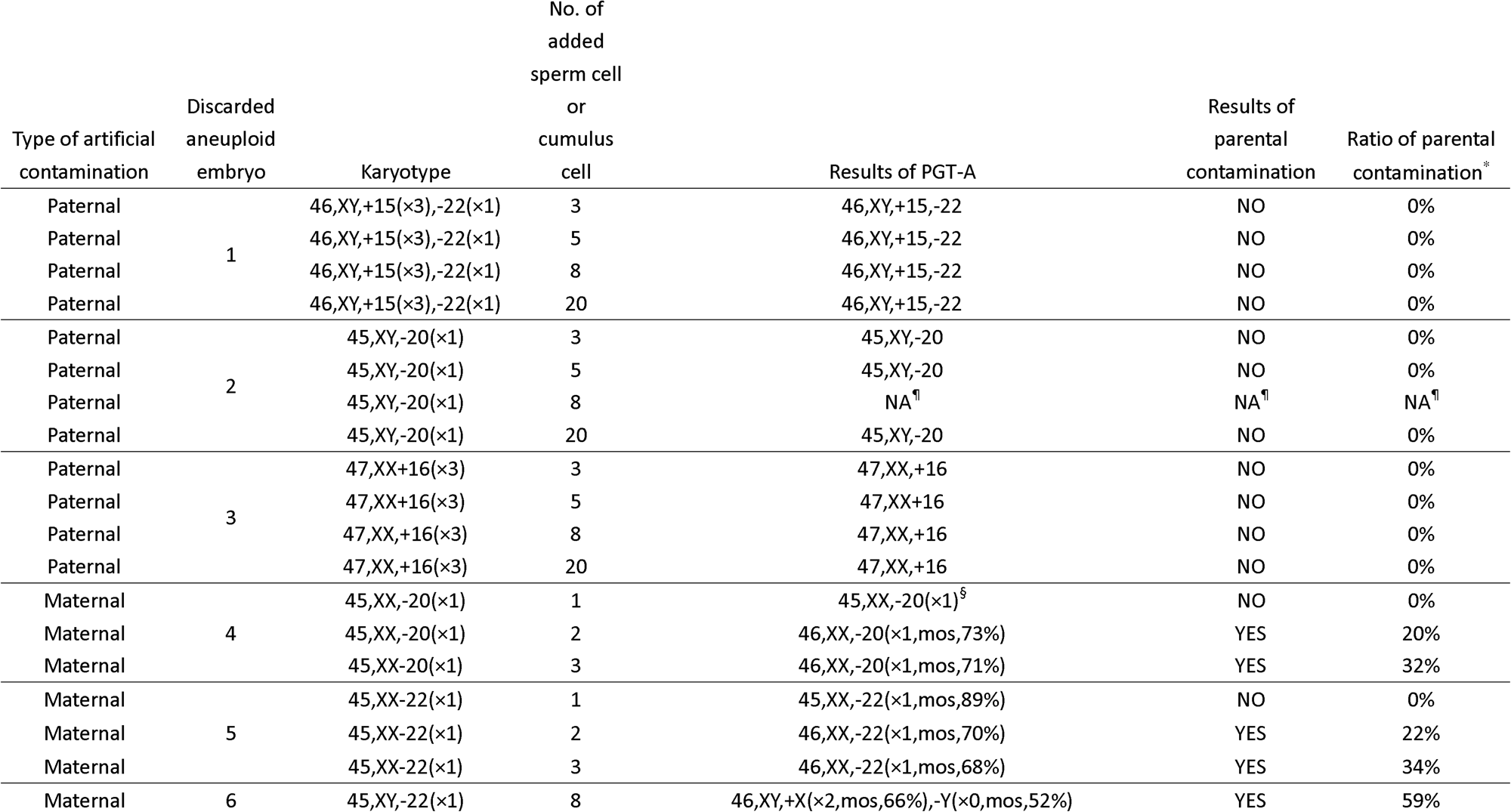
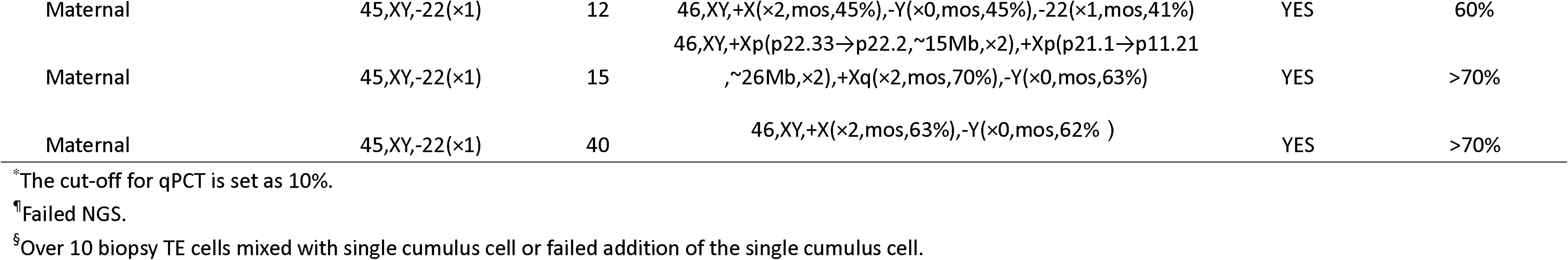
Validation results of the qPCT method in the artificial model.

To reveal the correlation between the PGT-A results and parental contamination, we selected discarded aneuploid blastocysts, added different numbers of sperm cells or cumulus cells to biopsied TE samples, and performed PGT-A and qPCT in these samples. The results showed that no paternal contamination was detected, and no change in the PGT-A results was observed, regardless of the number of added sperm cells (Figure 2A and Table 1). On the other hand, the proportion of maternal contamination increased gradually with the increase in the number of added cumulus cells, and a notable correlation was observed (Figure 2B and Table 1).

**Figure 2.**
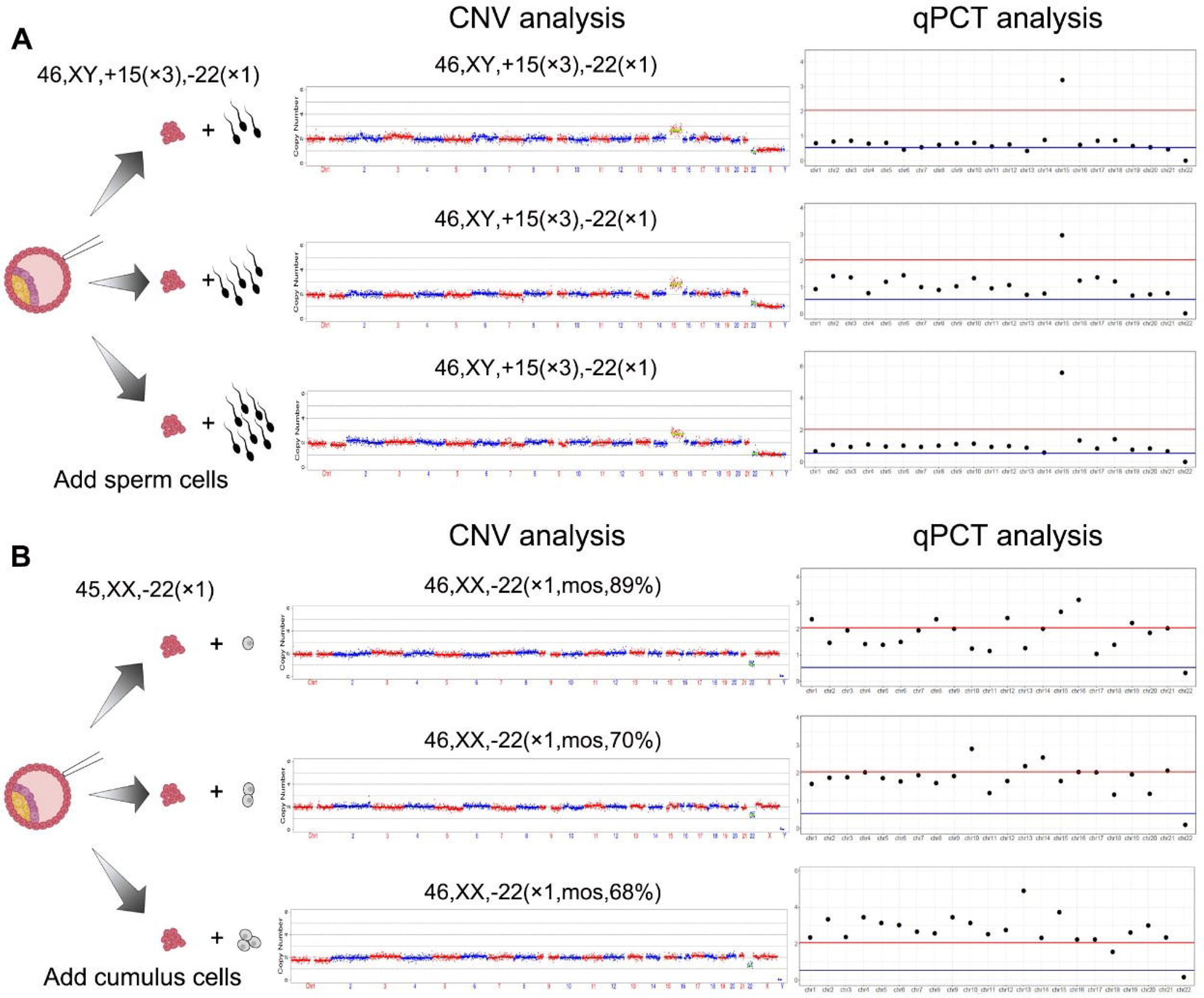
Validation of qPCT in the artificial model. **A:** Sperm cells (3 to 8 cells) were artificially added to approximately 10 biopsy TE cells respectively. **B:** Cumulus cells (1 to 3 cells) were artificially added to approximately 10 biopsy TE cells respectively.

### Prospective clinical study of qPCT-PGT-A in thawed IVF blastocysts

After the systematic validation of the qPCT assay, we performed qPCT-PGT**-**A in a total of 120 frozen blastocysts from 34 recruited patients as described in the materials. The procedure of qPCT-combined PGT-A is shown in (Figure. 3A) The qPCT results revealed a maternal contamination rate of 0.83% (1/120, case qpcts8), and no paternal contamination was detected, as expected (Table 2 and Figure 3B). Notably, one sample from case qpcts26 was defined as aneuploid with suspicious paternal contamination (Table 2 and Supplementary Figure S4A). To confirm this, the sample was collected and lysed for heteroploid detection using the whole embryo, and a paternal origin of triploid result was revealed (Table 2 and Supplementary, Figure S4B). The PGT**-**A results showed that the euploidy rate of these frozen blastocysts was 47.50% (57/120), and at least one euploid blastocyst was obtained for frozen embryo transfer (FET) in 28 of these patients (Table 2). Single-blastocyst transfer was performed in ternty-one patients, resulting in four healthy birth and ten ongoing clinical pregnancies (Table 2). To date, no early abortion has occurred, and we continue to follow up the last ongoing pregnancy. The detailed clinical indications and outcomes are summarized in Table 2.

**Figure 3.**
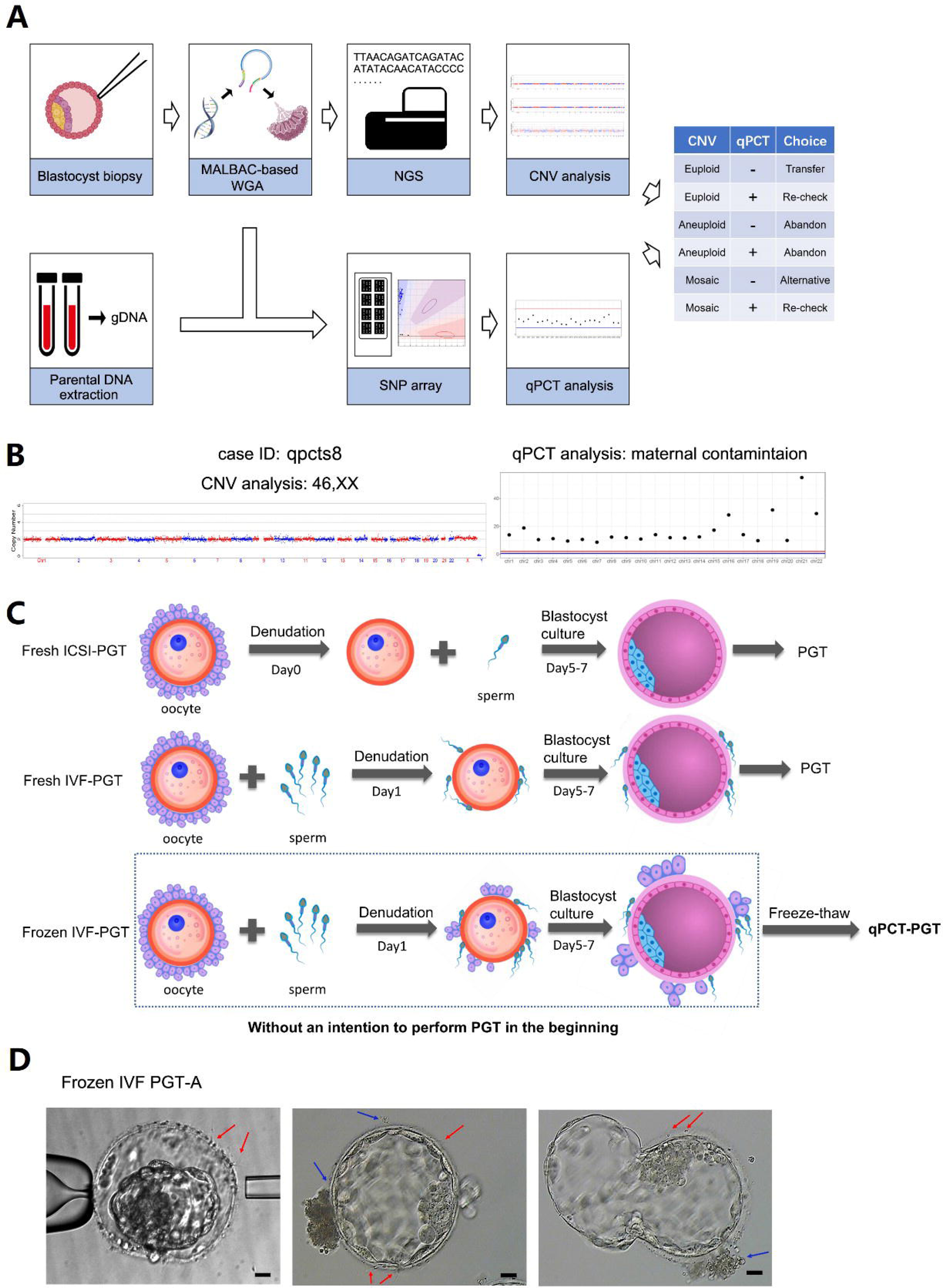
Clinical application of qPCT-combined PGT-A in human conventional IVF embryos. **A:** Workflow of qPCT-combined PGT-A. **B:** Clinical sample with positive results of qPCT. Case ID qpcts8: maternal contamination. **C:** Three clinical application scenarios of PGT. Fresh ICSI-PGT: PGT in fresh blastocysts from ICSI cycles. Fresh IVF-PGT: PGT in fresh blastocysts from conventional IVF cycles. Frozen IVF-PGT: PGT in frozen-thaw blastocysts from conventional IVF cycles. **D:** Typical IVF embryos undergoing TE cells biopsy. Bar = 10μm.

## Discussion

### WGA of sperm cells and the effect on PGT-A

Sperm are the main source of paternal contamination, and sperm attached to the zona pellucida cannot be removed in fresh or frozen embryos from conventional IVF cycles. Several studies have shown that sperm DNA is difficult to be amplified by using routine protocols for TE biopsy samples (15, 21), but little evidence indicating the risk and ratio of paternal contamination in conventional IVF PGT-A has been reported. In this study, a comprehensive investigation of paternal contamination was performed with a 3-step study design. First, we performed MALBAC-WGA of isolated sperm cells, and the results of both agarose gel electrophoresis and CNV-seq showed amplification failure, even if 20 sperm cells were tubed (Figure 1C). Although the immobilized motile sperm were subjected to freeze-thawing, the sperm remained intact and did not show signs of degeneration. Also, a similar negative result was found by Neelke et al using PicoPlex WGA (15). In contrast, sperm DNA could be amplified by using MDA (Figure 1C and 1E), indicating a higher risk of paternal contamination in the application of MDA-based PGT in IVF embryos. Second, we performed PGT-A and qPCT with artificially mixed TE and sperm cells, resulting in no change in the PGT-A results and no paternal contamination. Finally, we conducted a prospective clinical study with 120 frozen conventional IVF blastocysts; unsurprisingly, no paternal contamination was detected (Table 2). This evidence indicates that the risk of paternal contamination is negligible and that PGT can be applied in patients undergoing conventional IVF with the MALBAC system. As mentioned above, it is difficult to eliminate the possibility that sperm cells could spontaneously disruption in the real world or use other WGA systems.

### Maternal contamination from cumulus cells and the effect on PGT-A

Cumulus cells are the main source of maternal genetic contamination in TE samples. Unlike sperm, cumulus cell DNA can be easily amplified during the PGT procedure. A notable correlation between the number of cumulus cells and the proportion of maternal contamination was observed by using the qPCT method (Table 1). The results suggested that severe maternal contamination would lead to false-negative results, and partial maternal contamination would cause to mosaicism Therefore, to identify false-negative results caused by maternal contamination, it is necessary to apply contamination quantification methods, such as qPCT, in the PGT-A of both ICSI and conventional IVF embryos.

### Advantages and limitations of the qPCT method

In the PGT scenario, biopsy specimens from blastocysts usually require single-cell whole genome amplification to provide sufficient DNA for genetic testing. However, ADO (PCR failure for one allele) and/or preferential amplification (hypoamplification of one allele) resulting from WGA technology cause SNP genotyping to be unreliable (20). Therefore, many proposed parental cell contamination testing methods involving gDNA samples from tissue without WGA cannot be employed in PGT applications (17-19). For this reason, we developed a parental orientation test based on the allelic ratio determined via B allele frequency (BAF) analysis to address the parental contamination issue in IVF-PGT-A. Our method fully considers amplification biases, such as preferential amplification or ADO, which are relatively insensitive to single-cell WGA quality. Meanwhile, combined with WGA specific BAF pattern resolution, parental contamination or heteroploidy may be further discriminative (Supplementary Figure S4). Technically, any WGA method is applicable for qPCT. Also, preimplantation genetic testing for monogenic (PGT-M) as well as preimplantation genetic testing for chromosomal structural rearrangements(PGT-SR)are theoretically feasible for conventional IVF insemination method by using qPCT. The method does not require specialized equipment or complex experimental procedures; therefore, it can be fully adapted for routine use in a molecular diagnostic laboratory. However, this approach requires parental genetic information for combined analysis, thus limiting its large-scale clinical application to some degree. Moreover, a method of parental contamination removal is urgently needed to provide accurate PGT results for biopsy samples, even samples contaminated by parental genetic material. Epigenetic signal discrimination between embryonic and parental cell DNA may be a promising way to trace the cellular origin of DNA samples, which might filter out the effect of parental contamination in the future (22).

### The need for qPCT clinical application in frozen blastocysts

The biopsy of the existing frozen IVF blastocysts to meet patients’ clinical needs not only increases embryo utilization but also reduces physical and economic burdens. Thus, IVF physicians and embryologists must understand and select the programs available to them. The only barrier to this strategy is that the potential risk of parental DNA contamination cannot be excluded. Unlike ICSI-PGT-A or fresh IVF-PGT-A (17-19), cumulus cells are not required to completely removed in conventional IVF when there is no PGT expectation in advance (Figure 3C). As a result, it is difficult to always avoid sperm and/or cumulus cells attached to the spherical zona pellucida during TE cells biopsy (Figure 3D). Moreover, the DNA of sperm or granular cells disrupted by a laser might disperse into the biopsy fluid during biopsy. Therefore, it is necessary to detect parental contamination in biopsy cells under such conditions. Frozen IVF blastocyst biopsy samples will be resuscitated in advance, so the blastocyst TE cells will hatch from the holes generated by freezing and draining water or artificial openings made after thawing. In this study, 71.67% of blastocysts were stage 5 and 6 blastocysts with TE cells located far away from the zona pellucida (Supplementary Table 1), which could partly explain the reason for the low contamination detection rate in this study. Admittedly, the major disadvantages of the applied clinical strategy are that (□) it requires invasive embryo biopsy and (□) repeated cryopreservation of embryos is usually unavoidable. There are still safety concerns with these procedures (23-28). In the future, rapid noninvasive chromosome screening or mutation identification combined with qPCT will be a promising research area (29-33). However, as the clinical strategy yielded a clinical pregnancy rate of 57.14% without adverse outcomes, patients with existing frozen conventional IVF embryos could be counseled regarding this new option.

In summary, the risk of PGT in embryos with potential parental contamination is relatively low. We have established a practical and easy-to-implement clinical strategy, qPCT, for detecting potential parental contamination from sperm and cumulus cells especially in frozen conventional IVF embryos and ensuring the accuracy of PGT results. Our validation data and initial clinical applications strongly suggest that the qPCT-PGT assay can help to improve clinical outcomes via the chromosome screening of frozen conventional IVF embryos in a manner allowing their reuse. We envision that multicenter randomized clinical trials will be designed and performed in the future, which will further evaluate the clinical safety and effectiveness of the qPCT-PGT assay.

## Data Availability

All data produced in the present study are available upon reasonable request to the authors.

## Acknowledgements

We thank the patients for their participation in this study. This work was supported by grants from the Science and Technology Program of Guangzhou (201704020217), the National Key Research and Development Program of China (2018YFC1002604) and the National Natural Science Foundation of China (81800184).

## Author Contributions

Y.D., D.L., C.W., Y.Z., S.L., X.Z., F.L. designed research; Y.D., D.L., C.C., M.D., Y.H., C.H., H.W., X.Z., F.W., X.Z., N.L. performed research; Y.D., D.L., C.W., Y.Z., S.J., S.L., F.L. analyzed data; Y.D., D.L., C.W., Y.Z. wrote the paper.

