## Supplementary for "Evaluate the risk in conventional IVF frozen human blastocysts undergoing PGT using a new quantification method for parental contamination testing (qPCT)"

**Supplemental Figure 1.** Workflow of the simulate freezing procedure of sperm using immature oocytes zona pellucida. Motile sperm were incubated with four randomly selected MI or GV oocytes in 4-well culture plates for 2 h at 37°C under 6% CO2. After 2 h of incubation, the oocytes were transferred to G-MOPS-PLUS and were then flushed several times to dislodge loosely adherent sperm using a pipette approximately twice the diameter of the oocyte (250 mm) in three separate wells. The vitrification of the blastocysts was subsequently simulated. Thereafter, the sperm were blown off with a 120 micron stripping needle. A small aliquot was placed in a Petri dish and processed with a micromanipulator used for ICSI. Individual immobile dead sperm were aspirated and placed in PBS and tubed under the same conditions as the TE samples. There was no simulated vitrification procedure for the acquisition of fresh sperm. Cumulus cells were used to simulate the vitrification process; hyaluronidase digestion was performed to obtain single cells, which were then washed twice in G-MOPS-PLUS and processed with a biopsy needle in the same manner as TE samples.

**Supplemental Figure 2.** Distribution of relative POB statistics established by 30 reference samples from euploid embryos without parental contamination. (A) Chromosome level. (B) Genome level.

**Supplemental Figure 3.** Results of qPCT for parental DNA contaminated WGA products. (A) 30 ng MALBAC-WGA product of trophectoderm cells contaminated by 10 ng paternal DNA. (B) 30 ng MALBAC-WGA product of trophectoderm cells contaminated by 10 ng maternal DNA.

**Supplemental Figure 4.** Suspected parental contamination identification and heteroploidy detection for the embryo (case qpcts26). (A) The $R_{POB}$ of each chromosome indicates a paternal contamination or possible heteroploid. (B) A positive result of triploid was revealed according to the BAF pattern of filtered SNP loci. During the stage of prospective study of PGT-A in thawed IVF-blastocysts, we identified one sample (case qpcts26) as aneuploid with suspicious paternally genetic orientation (Figure S3A). To further confirm the result, the whole embryo was collected and amplified using MALBAC-based WGA. Again, genotyping for the amplified DNA was performed using the Infinium ASA bead chip as described above. SNP loci were filtered based on MALBAC amplification efficiency (lab study data), the triploid was then defined by using the BAF pattern of filtered SNP loci with four clusters of 0, 0.33, 0.67 and 1 (Figure S3B). Combined with these two analyses, paternal origin of triploid was then determined.

30 **Supplemental Figure 1**

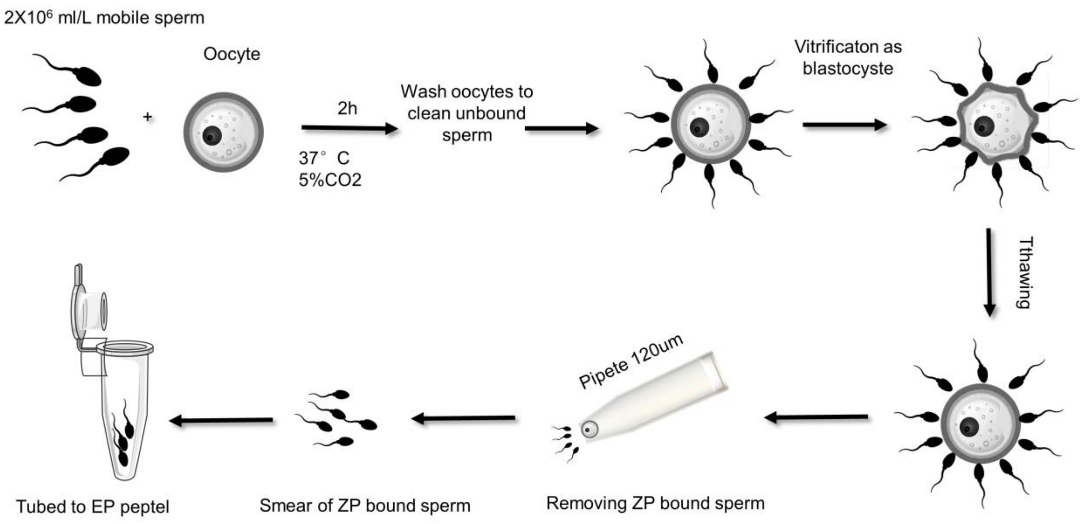

**Supplemental Figure2**

A

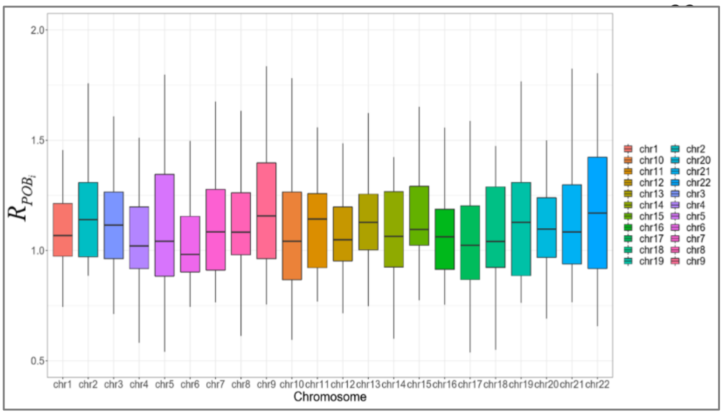


B

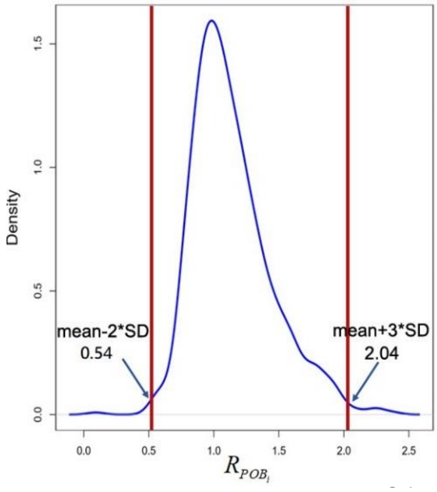

**Supplemental Figure3**

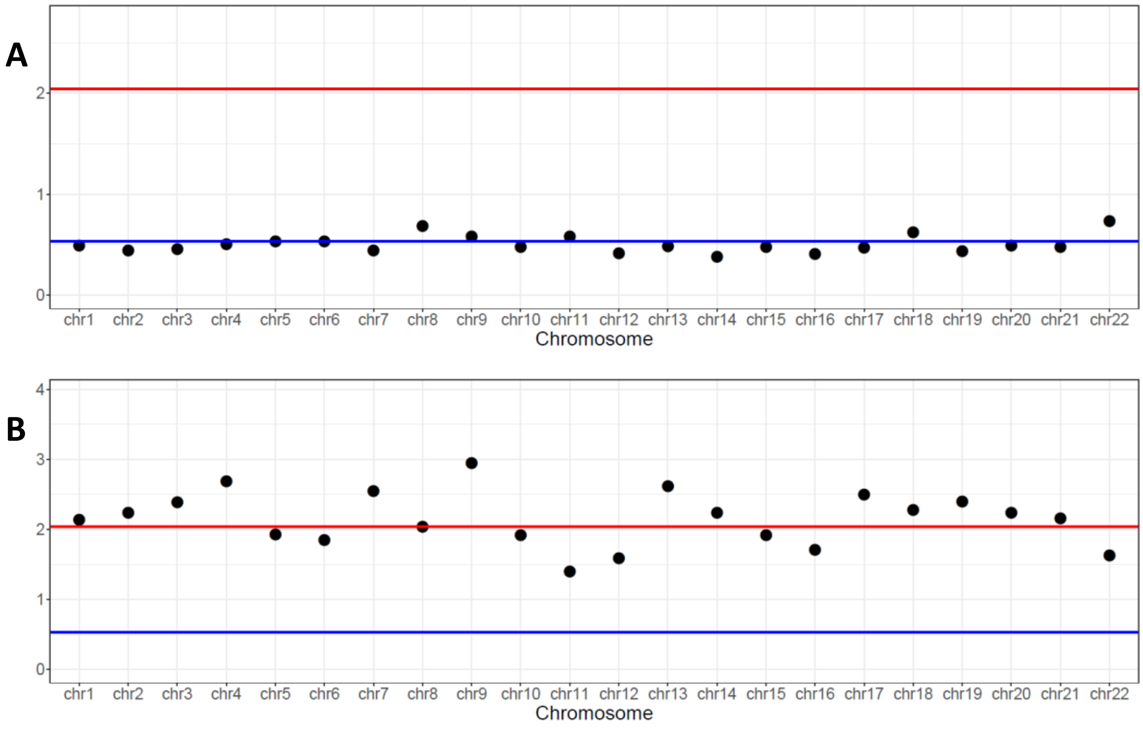

**Supplemental Figure 4**

**A**

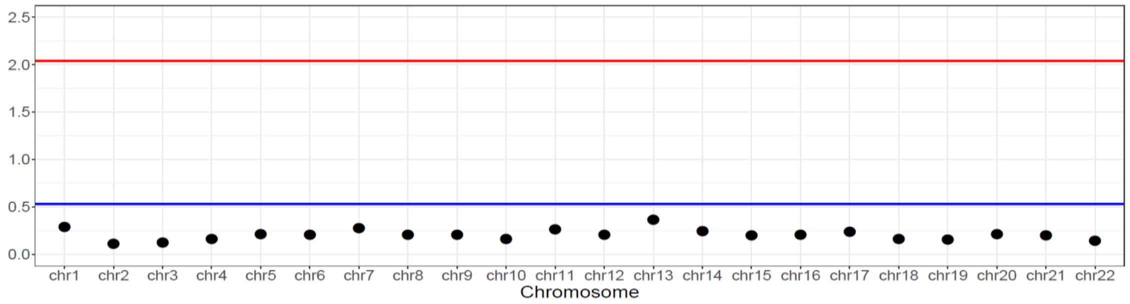


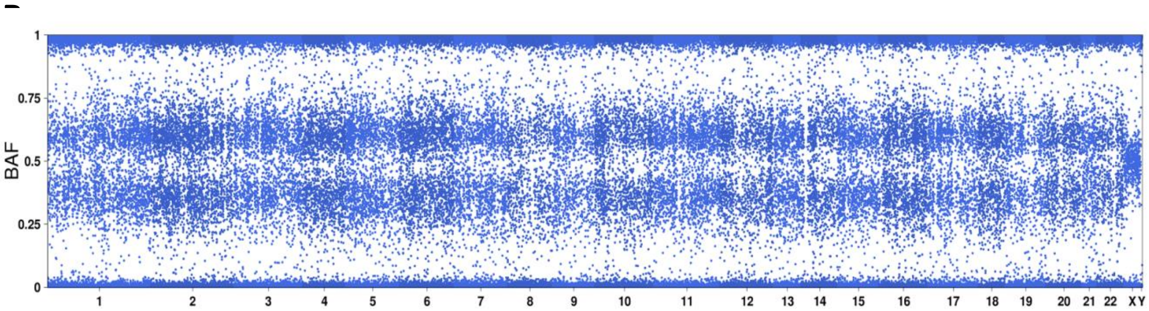

**Supplemental Table1. Summary of clinical information of the 34 patients subjected to PGT-A with qPCT.**

| Patient ID | Female age | PGT-A indication after previous IVF-ET | Total no. embryos tested | 4/5/6 stage blastocyst | Euploid embryos determined by PGT-A | qPCT assay | Frozen embryos transferred | Pregnancy outcome after FET | NIPT |
|---|---|---|---|---|---|---|---|---|---|
| qpcts1 | 31-35 | Abnormal pregnancy-labor history | 8 | 4/2/2 | 5 | Negative | 1 | Ongoing pregnancy | |
| qpcts2 | 36-40 | Abnormal pregnancy-labor history | 2 | 0/1/1 | 1 | Negative | 1 | Negative | |
| qpcts3 | 31-35 | Repeated implantation failure | 2 | 1/1/0 | 1 | Negative | 1 | Biochemical | |
| qpcts4 | 26-30 | Abnormal pregnancy-labor history | 5 | 0/2/3 | 5 | Negative | 1 | Live birth | Normal |
| qpcts5 | 36-40 | Abnormal pregnancy-labor history | 5 | 3/1/1 | 3 | Negative | 1 | Live birth | Normal |
| qpcts6 | 41-45 | Repeated implantation failure | 2 | 2/0/0 | 1 | Negative | 1 | Live birth | Normal |
| qpcts7 | 41-45 | Repeated implantation failure | 1 | 0/1/0 | 1 | Negative | 1 | Biochemical | Normal |
| qpcts8 | 36-40 | Repeated implantation failure | 5 | 1/4/0 | 2 | Positive* | 1 | Ongoing pregnancy | Normal |
| qpcts9 | 36-40 | Abnormal pregnancy-labor history | 3 | 0/3/0 | 1 | Negative | | | |
| qpcts10 | 36-40 | Abnormal pregnancy-labor history | 2 | 1/1/0 | 0 | Negative | | | |
| qpcts11 | 31-35 | Repeated implantation failure | 4 | 0/2/2 | 2 | Negative | | | |
| qpcts12 | 36-40 | Abnormal pregnancy-labor history | 1 | 1/0/0 | 1 | Negative | 1 | Biochemical | |
| qpcts13 | 31-35 | Abnormal pregnancy-labor history | 2 | 0/2/0 | 2 | Negative | 1 | Ongoing pregnancy | Normal |
| qpcts14 | 41-45 | Repeated implantation failure | 5 | 0/5/0 | 1 | Negative | 1 | Negative | |
| qpcts15 | 31-35 | Abnormal pregnancy-labor history | 3 | 2/1/0 | 3 | Negative | 1 | Negative | |
| qpcts16 | 36-40 | Repeated implantation failure | 2 | 1/1/0 | 0 | Negative | | | |
| qpcts17 | 26-30 | Repeated implantation failure | 3 | 0/3/0 | 0 | Negative | | | |
| qpcts18 | 36-40 | Repeated implantation failure | 1 | 1/0/0 | 0 | Negative | | | |
| qpcts19 | 41-45 | Repeated implantation failure | 3 | 2/0/1 | 1 | Negative | | | |
| qpcts20 | 31-35 | Abnormal pregnancy-labor history | 5 | 1/4/0 | 3 | Negative | 1 | Ongoing pregnancy | Normal |
| qpcts21 | 41-45 | Abnormal pregnancy-labor history | 1 | 0/0/1 | 1 | Negative | 1 | Live birth | Normal |
| qpcts22 | 26-30 | Repeated implantation failure | 5 | 0/5/0 | 2 | Negative | | | |
| qpcts23 | 26-30 | Repeated implantation failure | 7 | 2/5/0 | 5 | Negative | | | |
| qpcts24 | 31-35 | Abnormal pregnancy-labor history | 2 | 0/2/0 | 0 | Negative | | | |
| qpcts25 | 31-35 | Abnormal pregnancy-labor history | 4 | 0/4/0 | 2 | Negative | 1 | Ongoing pregnancy | |

| | | | | | | | | |
|---|---|---|---|---|---|---|---|---|
| qpcts26 | 31-35 | Abnormal pregnancy-labor history | 5 | 1/4/0 | 1 | False-Positive§ | 1 | Ongoing pregnancy |
| qpcts27 | 36-40 | Abnormal pregnancy-labor history | 1 | 1/0/0 | 1 | Negative | 1 | Ongoing pregnancy |
| qpcts28 | 21-25 | Repeated implantation failure | 3 | 2/1/0 | 3 | Negative | | |
| qpcts29 | 36-40 | Repeated implantation failure | 5 | 0/5/0 | 1 | Negative | 1 | Ongoing pregnancy |
| qpcts30 | 36-40 | Abnormal pregnancy-labor history | 6 | 4/2/0 | 2 | Negative | 1 | Ongoing pregnancy |
| qpcts31 | 36-40 | Abnormal pregnancy-labor history | 3 | 2/1/0 | 0 | Negative | | |
| qpcts32 | 26-30 | Abnormal pregnancy-labor history | 3 | 2/0/1 | 2 | Negative | 1 | Ongoing pregnancy |
| qpcts33 | 31-35 | Abnormal pregnancy-labor history | 3 | 0/1/2 | 1 | Negative | | |
| qpcts34 | 41-45 | Repeated implantation failure | 8 | 0/8/0 | 3 | Negative | 1 | Negative |
| Total | | | 120 | 34/72/14 | 57 | 1 embryo | 21 | 14 Clinical pregnancies |

*One of five IVF embryos form case 182866 was detected as maternal contamination.

§A paternal origin of triploid.
